# Low-MBL producing genotypes playing a role on protection to leprosy and its multibacillary forms in a highly endemic area for leprosy in Brazil

**DOI:** 10.1101/2022.08.22.22279041

**Authors:** Erika M. de Salles, Lívia M. Martins, Rita de C. M. Escocard, Bruno V. G. Forte, Yonnara E. C. Maciel, Thereza L. Kipnis, Edilbert P. Nahn, Wilmar D. da Silva, Alba L. Peixoto-Rangel

**Affiliations:** Instituto de Ciências Biomédicas, Departamento de Imunologia, Universidade de São Paulo, São Paulo, SP, Brazil; Laboratório de Biologia do Reconhecer, Centro de Biociências e Biotecnologia, Universidade Estadual do Norte Fluminense Darcy Ribeiro, Campos dos Goytacazes, RJ, Brazil; Faculdade de Medicina de Campos, Campos dos Goytacazes, RJ, Brazil; Laboratório de Imunoquímica, Instituto Butantan, São Paulo, SP, Brazil

**Keywords:** Leprosy, Mannose binding lectin, Polymorphism, *Mycobaterium leprae*, SNV, Complement System

## Abstract

The high frequency of *MBL2* mutant alleles found in various human populations suggests that they provide some selective advantage. One of the hypotheses is that MBL deficiency may be protective against intracellular pathogens, including *Mycobacterium leprae*. In this study, we investigated the role of *MBL2* in the susceptibility to leprosy. Single-nucleotide variants (SNV) B (rs1800450) and C (rs1800451) in exon 1 of the *MBL2* gene and serum MBL concentrations were assessed in 53 patients with leprosy and 68 healthy controls from southeast Brazil by polymerase chain reaction followed by restriction fragment length polymorphism and commercial capture enzyme-linked immunosorbent assay, respectively. The genotyped SNVs were significantly associated with increased protection against leprosy (OR=0.3757, 95% CI=0.1615⍰08740, p=0.0273) and more severe forms, including multibacillary (OR=0.3769, 95% CI=0.1447⍰0.9818, p=0.0493) and lepromatous (OR=0.2423, 95% CI=0.06547⍰0.8968, p=0.0369) leprosy. On the other hand, low-MBL producing genotypes were associated with the protection for leprosy, especially in its more severe forms. Low MBL levels and low MBL producing variants were associated with protection against leprosy and progression to more severe forms of this disease. Moreover, this study highlights MBL as a regulator of immune function in which alterations may affect normal responses to leprosy infection and inflammatory stimuli.

## Introduction

Leprosy is a chronic infectious disease caused by the obligate intracellular bacillus *Mycobacterium leprae* that primarily affects the skin and peripheral nerves.^1^ Most individuals are inherently resistant to leprosy and only a small group develops the disease after exposure to *M. leprae* for long periods. Leprosy is characterized by a wide spectrum of immunologically-defined clinical manifestations. The most severe manifestation is the lepromatous form, characterized by the absence of specific cellular immunity leading to widely disseminated disease and extended multibacillary lesions. The other end of the spectrum is represented by the tuberculoid form, which is associated with a vigorous Th1 response and paucibacillary lesions.^2,3^ Innate immunity molecules such as mannose-binding lectin (MBL) play an important role on host defense against infections due to their ability to activate the complement system and mediate the lysis and opsonization of microorganisms, and may be involved in the modulation of the immune response to infection with *M. leprae*.^4^ The mannose-binding lectin (MBL) pathway involves a C-type lectin, which binds to mannose-containing carbohydrates in microorganisms. MBL levels are genetically determined and may be associated with increased susceptibility to infection by viruses, bacteria, fungi, and protozoa.^5, 6, 7^ However, it has been proposed that MBL deficiency may protect the host against intracellular entry by *M. leprae* by reducing the opsonization through the deposition of multiple C3b fragments on the microbial cell surface, resulting in lower phagocytic uptake of bacilli. ^8, 22^ MBL acts as a pattern recognition molecule for a wide range of infectious agents, recognizing sugar moieties such as mannose, N-acetylglucosamine, fucose, and glucose present on the surface of several microorganisms, leading to their phagocytosis and activation of complement.^9, 19^

In humans, the gene encoding *MBL2* is located on the long arm of chromosome 10 (q11.2-q21) and includes four exons. MBL serum levels are strongly affected by structural mutations in exon 1 and promoter region of the gene. A single-nucleotide variant (SNV) was found at codon 54 (allele B, Gly54Asp) in exon 1 of the *MBL2* gene,^10^ and subsequent studies have revealed the presence of two SNVs at codons 52 (allele D, Arg52Cys) and 57 (allele C, Gly57Glu).^11^ The alleles derived from SNVs at codons 52, 54, and 57 are termed *MBL-D, MBL-B*, and *MBL-C*, respectively, and are associated with a significant decrease in MBL levels as compared to levels in individuals carrying both copies of the normal allele, termed *MBL-A*.^11, 12, 13^ In addition, three SNVs (termed “H/L”, “Y/X”, and “P/Q”) in the promoter region affect MBL serum concentration. The combination of SNVs results in three haplotypes (HYPA, LYPA, and LYQA) that are associated with normal MBL levels and four haplotypes (LXPA, LYPB, LYQC, and HYPD) associated with low or very low MBL levels.^14^ It is possible that these variants have been maintained at high frequencies because they provide some selective advantage to the host.^9, 11, 15^ Thus, MBL variants may be conserved mostly in heterozygous individuals. Even though some studies have shown the involvement of the complement system in leprosy, few have investigated the role of *MBL2* gene variants on infection with *M. leprae* and disease progression. In addition, the genetic diversity of human populations hints at the existence of yet undescribed point mutations in some polymorphic genes that warrant investigation, particularly those with a putative involvement in infectious diseases. In Brazil, infection with *M. leprae* has been reported in different geographic regions.^21, 22, 24^ However, these findings have not been extensively validated. Here we investigated a geographically distinct population from the Southeast Brazil in order to evaluate the role of the *MBL2* gene and MBL protein in leprosy susceptibility. The present study provides novel information on the association between *MBL2* gene variant and susceptibility to leprosy across the spectrum of leprosy forms.

## Material and Methods

### Subjects

Cases and controls were recruited in Campos dos Goytacazes, Rio de Janeiro, southeast Brazil (21° 45’ 15” S and 41° 19’ 28” W, 13 m asl). Fifty-three patients with leprosy were selected at the Hansen Health Program of the Campos dos Goytacazes Health Secretariat, a regional reference center for treatment of leprosy. Diagnosis was based on clinical examination and bacilloscopy of suspected tissue lesions. Bacillary lesion counts and associated typical histopathology parameters permit diagnosis and classification of leprosy forms (Table 1).^16, 17^ Healthy controls consisted of 68 unrelated individuals recruited from the local blood bank (Table 1). Patients (L) and control subjects (HC) were from the same geographical area. Leprosy patients were grouped according to the World Health Organization (WHO) classification in multibacillary (MB) or paucibacillary (PB) leprosy and the Madrid classification in lepromatous leprosy (LL), dimorph leprosy (DL), indeterminate leprosy (IDL), and tuberculoid leprosy (TL) for the analysis (Table 1).The exclusion criteria for subjects were other infectious or inflammatory disease. Informed consent (written) was obtained from all participants and the study was approved by the local Medical Ethics Committee (n°. 1.687.476).

**Table 1:**
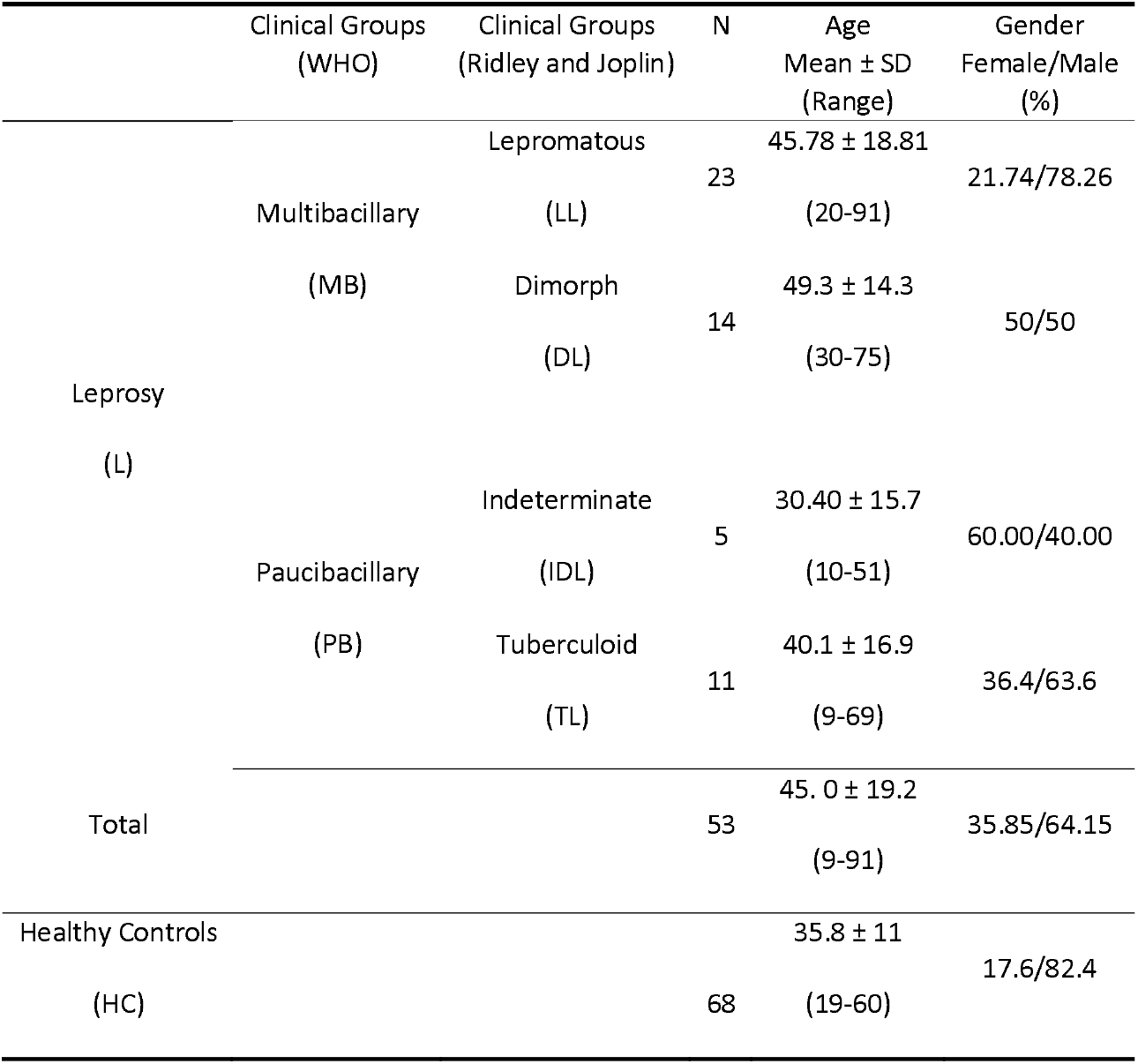
Clinical profile of patients with leprosy.

### Serum MBL level measurement

Serum MBL concentrations were determined using a commercial capture enzyme-linked immunosorbent assay (MBP-oligomer ELISA, Microwell strip version; Antibody Shop, Copenhagen, Denmark). Serum (1:100) was added to ELISA microplates coated with monoclonal antibody against the oligomeric MBL carbohydrate-binding domain. The bound MBL was detected by a second biotin-labeled antibody and streptavidin peroxidase, followed by the addition of tetramethylbenzidine (TMB). Color intensity was read in an ELISA reader at 450 nm (Multiskan EX Primary EIA, Thermo Fisher, Waltham, MA, USA). The serum standard results were used to build a calibration curve. MBL concentrations are defined as low (< 100 ng/ml), intermediate (100⍰1000 ng/ml), and high (< 1000 ng/ml) according to the kit manufacturer’s instructions.

### *MBL2* genotyping

The exon 1 region of *MBL2* (codons 54 and 57 for alleles B and C, respectively) was genotyped by allele-specific polymerase chain reaction followed by restriction fragment length polymorphism (PCR-RFLP). The forward and reverse primers used were 5’1 AGTCGACCCAGATTGTAGGACAGAG⍰3’ and R: 5’ ⍰AGGATCCAGGCAGTTTCCTCTGGAA GG13’,respectively, as described elsewhere.^18^ The PCR was performed in 25-μl volumes containing 1001500 ng (2 μl) of genomic DNA, 0.5 μl of specific primers (12.5 pmol/ml; Fermentas, Waltham, MA, USA), 10 mM deoxynucleotide triphosphates (Fermentas, USA), 1.25 μl MgCl₂ (50 mM; Invitrogen, Carlsbad, CA, USA), 2.5 μl 10× PCR buffer (without magnesium), and 1 U of *taq* platinum polymerase (Invitrogen, USA). The following reaction conditions were used: 94 °C for 2 min, 35 cycles at 94 °C for 30 s, followed by annealing at 60 °C for 30 s and 72 °C for 2 min, and a final extension step of 5 min at 72 °C. The PCR products were identified by electrophoresis in 2% (w/v) agarose gel stained with ethidium bromide. The fragment amplified by PCR (349 bp) was digested using *Ban*I and *Mbo*II restriction enzymes to detect B and C alleles. The *Ban*I restriction enzyme cleaved the MBL*A allele into two fragments (260 and 89 bp) and the MBL*B allele was not cleaved; *Mbo*II cleaved the MBL*C allele into two fragments (279 and 70 bp). Fragments were visualized by electrophoresis in 8% polyacrylamide gel stained with ethidium bromide.

### Statistical analysis

MBL levels were compared between leprosy patients and healthy controls using the Kruskal-Wallis test followed by Dunn’s or Mann-Whitney tests. The allelic and genotypic frequencies were compared using Fisher’s exact test. The associations were represented by odds ratios (ORs) with 95% confidence intervals (CIs). Differences were considered significant at p < 0.05. The analyses were performed using GraphPad Prism™ 5.0 (GraphPad Software, San Diego, CA, USA). Hardy-Weinberg equilibrium (HWE) was calculated for each SNV in a set of unrelated controls with a customized spreadsheet in Microsoft Excel.

## Results

### Serum MBL levels in patients and healthy controls

MBL serum levels ranged from 0.0 to 3903.0 ng/ml (median = 1983.0 ng/ml) in HC and 0.1 to 3024.8 ng/ml (median = 560.0 ng/ml) in L patients. There were no significant differences between L and HC groups (p = 0.2042) and in the comparison among patients with severe forms of the disease (MB and LL) and patients with less severe forms (p= 0.3588 and 0.5678, respectively) (Fig. 1). Leprosy patients were also assessed according to MBL production levels. The frequency of low-(< 100 ng/ml), intermediate-(100⍰1000 ng/ml), and high-(> 1000 ng/ml) MBL producers was similar between groups with different degrees of disease severity (Fig. 1).

**Figure 1:**
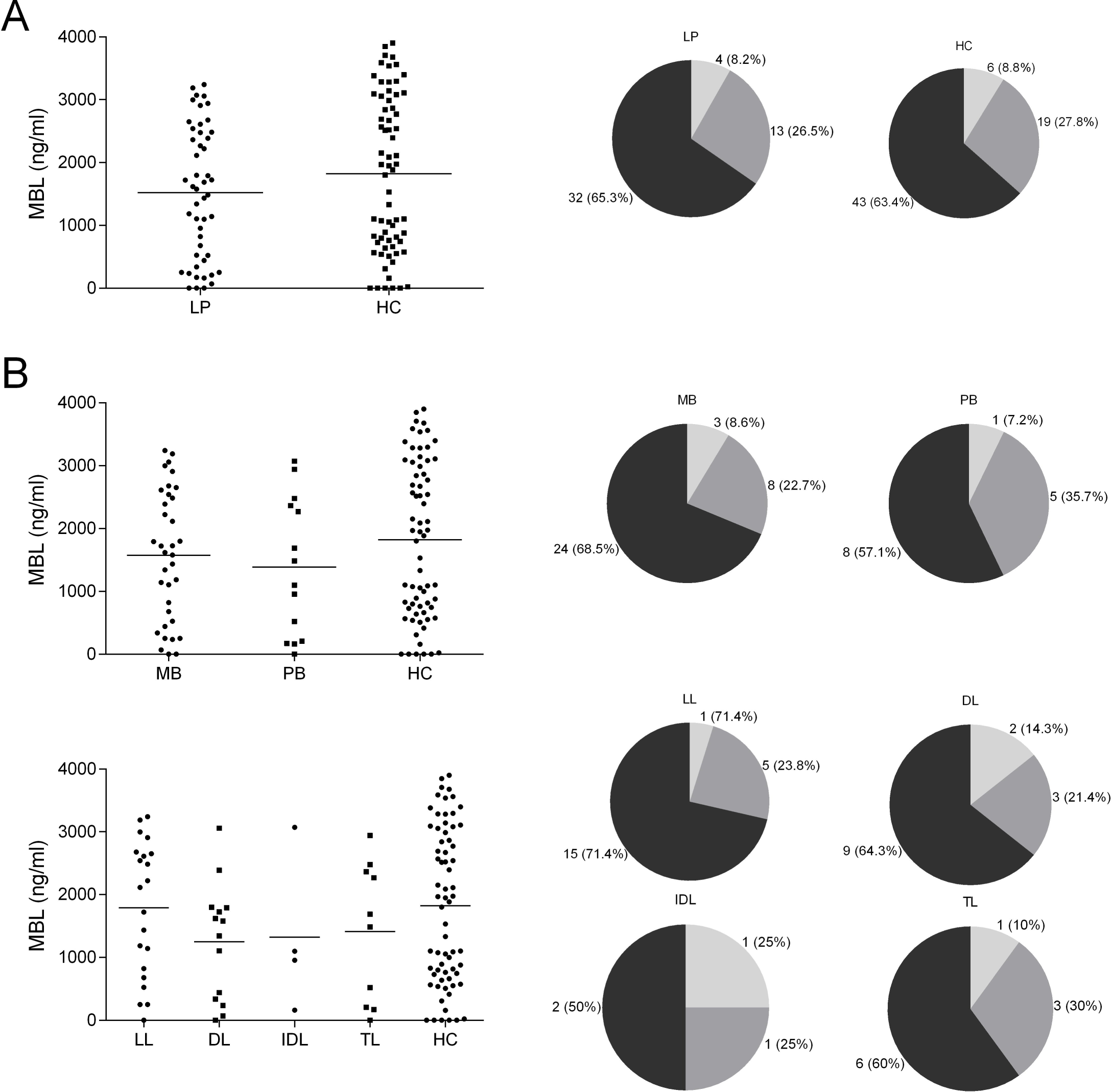
Serum MBL levels (dot plot graphs), and frequencies (parts of whole graphs) were measured by ELISA and plotted according to the presence or absence of leprosy (**A**); or according to WHO classification (**B**) or Madri classification (**C**). Individuals were considered low producers with up to 100 ng/ml, medium with 100 to 1000 ng/ml and high producers above 1000 ng/ml of MBL in sera.

**Figure 2:**
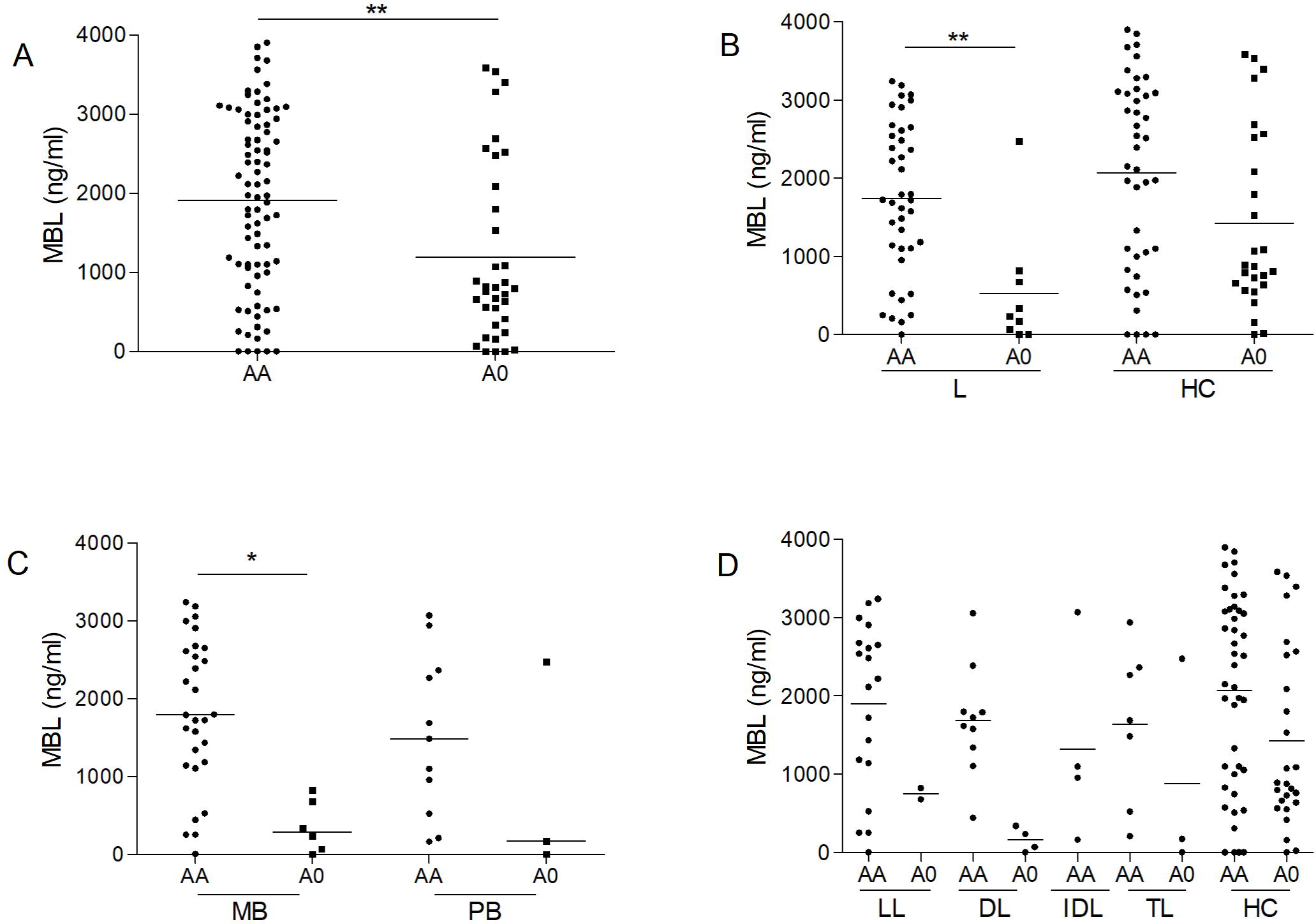
MBL production associated with *MBL2* genotypes (A) in all subjects and (B) in leprosy patients and healthy control. The levels of MBL were compared with Mann-Whitney test. Significance was considered at p < 0.05, p < 0.01 or p < 0.001 as represented by *, ** and ***, respectively.

### Allelic and genotypic distribution of SNV B (rs1800450) and C (rs1800451)

Only three individuals in the studied population had the C (rs1800451) allele: one individual with the DL form and two HCs, corresponding to a frequency of 0.025. The B allele (rs1800450) had a frequency of 0.2 in the population. When grouped according to presence or absence of leprosy, the frequencies of alleles B and C in the L and HC groups were 0.08 and 0.17, respectively. The presence of allele B was protective for the development of leprosy (OR = 0.375, 95% CI = 0.1567⍰0.8977, p = 0.0387) and the progression to multibacillary forms of the disease (OR = 0.348, 95% CI = 0.1298⍰0.9704, p = 0.0441) (**Table 2**).

**Table 2:** Allele and genotype distribution in subjects with leprosy and healthy controls relative to the single nucleotide variants B.

| Clinical form | Genotype AA<br>(frequency) | Genotype AB<br>(frequency) | *p* | OR<br>(95% CI) | Allele A<br>(frequency) | Allele B<br>(frequency) | *p* | OR<br>(95% CI) |
| --- | --- | --- | --- | --- | --- | --- | --- | --- |
| **Lepromatous** | 20<br>(0.87) | 3<br>(0.13) | 0.0633 | 0.275<br>(0.07407 - 1.021) | 43<br>(0.935) | 3<br>(0.065) | 0.0915 | 0.3256<br>(0.09319 - 1.138) |
| **Borderline** | 11<br>(0.786) | 3<br>(0.214) | 0.3691 | 0.5500<br>(0.1379 - 2.193) | 25<br>(0.893) | 3<br>(0.107) | 0.5755 | 0.5600<br>(0.1563 - 2.007) |
| **Indeterminate** | 5<br>(1) | 0<br>(0) | 0.1644 | 0.1651<br>(0.008751 - 3.116) | 10<br>(1) | 0<br>(0) | 0.3689 | 0.2187<br>(0.1238 - 3.861) |
| **Tuberculoid** | 8<br>(0.727) | 3<br>(0.273) | 0.7402 | 0.6875<br>(0.1666 - 2.837) | 19<br>(0.864) | 3<br>(0.136) | 0.7690 | 0.7368<br>(0.2018 - 2.691) |
| **Paucibacilar** | 13<br>(0.813) | 3<br>(0.227) | 0.2472 | 0.4231<br>(0.1096 - 1.163) | 29<br>(0.906) | 3<br>(0.094) | 0.2989 | 0.4828<br>(0.1358 - 1.716) |
| **Multibacilar** | 31<br>(0.838) | 6<br>(0.162) | **0.0441** | **0.3548**<br>**(0.1298 - 0.9704)** | 68<br>(0.919) | 6<br>(0.081) | 0.0654 | 0.4118<br>(0.1602 - 1.059) |
| **Leprosy** | 44<br>(0.83) | 9<br>(0.17) | **0.0387** | **0.3750**<br>**(0.1567-0.8977)** | 97<br>(0.915) | 9<br>(0.085) | 0.0578 | **0.4330**<br>**(0.1920 - 0.9763)** |
| **Control** | 44<br>(0.647) | 24<br>(0.353) | - | - | 112<br>(0.824) | 24<br>(0.176) | - | - |

To evaluate the protective role of variants in the *MBL* gene in the development and severity of leprosy, the frequency of alleles B and C was combined in a group named A0. Overall, *MBL* variants were associated with increased protection against leprosy (OR = 0.3757, 95% CI = 0.1615⍰0.8740, p = 0.0273), and their most severe forms according to WHO (MB: OR = 0.3769, 95% CI = 0.1447⍰0.9818, p = 0.0493) and Madrid classifications (LL: OR = 0.2423, 95% CI = 0.06547⍰0.8968, p = 0.0369) (**Table 3**). These results show that variants B and C are protective against the development of leprosy, especially in its more severe forms.

**Table 3:** Allele and genotype distribution in subjects with leprosy and healthy controls relative to the single nucleotide variants B and C combined.

| Clinical form | Genotype AA<br>(frequency) | Genotype AO<br>(frequency) | *p* | OR<br>(95% CI) | Allele A<br>(frequency) | Allele O<br>(frequency) | *p* | OR<br>(95% CI) |
| --- | --- | --- | --- | --- | --- | --- | --- | --- |
| **Lepromatous** | 20<br>(0.87) | 3<br>(0.13) | **0.0369** | **0.2423**<br>**(0.06547 – 0.8968)** | 43<br>(0.935) | 3<br>(0.065) | 0.0603 | 0.2952<br>(0.08488 – 1.026) |
| **Borderline** | 10<br>(0.714) | 4<br>(0.286) | 0.5583 | 0.6462<br>(0.1834 - 2.275) | 24<br>(0.857) | 4<br>(0.143) | 0.7885 | 0.7051<br>(0.2251 – 2.209) |
| **Indeterminate** | 5<br>(1) | 0<br>(0) | 0.1533 | 0.1458<br>(0.007737 – 2.747) | 10<br>(1) | 0<br>(0) | 0.2094 | 0.1986<br>(0.1127 – 3.499) |
| **Tuberculoid** | 8<br>(0.727) | 3<br>(0.273) | 0.7375 | 0.6058<br>(0.1472 – 2.492) | 19<br>(0.864) | 3<br>(0.136) | 0.7677 | 0.6680<br>(0.1837 - 2.429) |
| **Paucibacilar** | 13<br>(0.812) | 3<br>(0.188) | 0.2415 | 0.3728<br>(0.09687 – 1.435) | 29<br>(0.906) | 3<br>(0.094) | 0.2972 | 0.4377<br>(0.1237 - 1.548) |
| **Multibacilar** | 30<br>(0.811) | 7<br>(0.189) | **0.0493** | **0.3769**<br>**(0.1447 - 0.9818)** | 67<br>(0.905) | 7<br>(0.095) | 0.0757 | 0.4420<br>(0.1818 – 1.075) |
| **Leprosy** | 43<br>(0.811) | 10<br>(0.189) | **0.0273** | **0.3757**<br>**(0.1615 – 0.8740)** | 96<br>(0.906) | 10<br>(0.094) | **0.0448** | **0.4407**<br>**(0.2022 - 0.9606)** |
| **Control** | 42<br>(0.727) | 26<br>(0.273) | - | - | 110<br>(0.809) | 26<br>(0.191) | - | - |

### MBL serum levels and their association with MBL genetic variants

MBL serum levels were significantly lower in genotype A0 individuals than in AA individuals (p = 0.0025). When comparing A0 and AA genotypes in L and HC subjects, L individuals with A0 genotype had significantly lower MBL serum levels than those with AA genotype (p=0,0021). Thus, the results indicate that carrying the ancestral genotype (AA) is related to the highest levels of MBL, while to have the polymorphic variant (A0) can be associated to lower MBL production.

## Discussion

In this study, we investigated a geographically distinct population from the Southeast Brazil in order to evaluate the role of *MBL2* gene and MBL protein in the leprosy susceptibility. We found no significant association between MBL levels and leprosy when the analysis did not separate individuals by their genotypes, i.e, considering only the levels of MBL in the patients and controls sera (Fig.1). The normal levels of MBL in serum are very stable^25^, but may range from 3 to 50 ng/ml in different individuals. Due to this wide variation in MBL levels and since sample size was very small, this dosage shall be further tested in larger samples.

The variation in MBL levels can be explained by the occurrence of four allotypes due to mutations in exon 1 of the gene encoding the MBL polypeptide as well as several variants in the promoter region. In our study, we evaluated the genetic variants in exon 1 of *MBL2* gene (B and C alleles) and found that B variant (rs1800450) was associated with increased protection against leprosy (OR = 0.375, 95% CI = 0.1567⍰0.8977, p = 0.0387) and multibacillary forms of this disease (OR = 0.348, 95% CI = 0.1298⍰0.9704, p = 0.0441), and this effect remained significant even after alleles B and C were combined in a group called A0 (leprosy: OR = 0.3757, p = 0.0273; multibacillary form, MB: OR = 0.3769, p = 0.0493; and lepromatous leprosy, LL: OR = 0.2423, p = 0.0369). Our findings are partially similar to those of Sapkota et al.,^22^ who suggested a protective role for MBL deficiency against the development of the more severe multibacillary form of leprosy, but not for leprosy. However, their findings were based on protein levels only, unlike the our study that combined data on MBL levels and *MBL2* variant to define MBL deficiency. Another interesting finding of the current study is the distribution of *MBL2* genotypes (AA and A0): the frequencies of genotypes associated with low and intermediate MBL levels were significantly higher in leprosy (L) patients with A0 genotype than with AA genotype (p = 0.0021). This result indicates that genotype A0 was associated with significantly decreased risk of leprosy and manifestation of more severe forms of the disease, lending further support to the idea that MBL deficiency is associated with protection against leprosy and the development of multibacillary disease. Pathogen or damage-associated molecular sugar patterns (PAMPs/DAMps) on *M. leprae* are recognized by mannose-binding lectin (MBL) and activate the lectin pathway of complement. Autoactivation of the serine protease MASP-1 is followed by transactivation of MASP-2, cleavage of C2 and C4 and formation of the C3 convertase C2Ac4B. C3 convertase ultimately leads to the insertion of C3b opsonins on the surface of the pathogen (20). Also, increased MBL protein in serum is expected to enhance the uptake of C3b/C4b/ficolin/MBL-opsonized *M. leprae*. Thus, MBL genetic variants *rs1800450* and *rs1800451* herein associated to low MBL levels, may be contributing to reduce complement activation of the lectin pathway and phagocytosis, which could be highly beneficial, protecting against leprosy disease. However, functional investigations on phagocytic efficiency of the associated genetic variants *rs1800450* and *rs1800451* would help to clarify the role of this protein in the disease. Later, yet unidentified intrinsic pathways protect *M. leprae* bacilli from macrophage defenses by inhibiting the C5b-C9 membrane lytic complex^20^. Nevertheless, the molecular mechanisms of immune evasion by *M. leprae* bacilli must be better understood.

Messias-Reason et al. ^20^ showed for the first time an association between low-MBL producing haplotypes/compound genotypes and protection against development of lepromatous and borderline leprosy. However, they also found a significant association between the high-MBL producer haplotype LYPA and both leprosy per se and its lepromatous and borderline forms, a finding similar to those described in our study.

## Conclusions

In conclusion, this study supports a pivotal role for the MBL pathway in the susceptibility to leprosy. We showed that MBL genetic variants modulate the immune response in *M. leprae*-affected populations and are probably maintained by balancing selection, supporting previous studies of MBL and leprosy. ^21^ MBL seems to play an important role on the immune response to infection with *M. leprae*: low MBL levels and low MBL producing variants were associated with protection against leprosy and progression to more severe forms of this disease.

Considering the high efficiency of the molecular procedure used to identify MBL haplotypes, further studies are needed to clarify the role of MBL in the fate of infected cells. Investigation involving the role of pattern recognition receptors in recognizing pathogen-associated molecular patterns, certainly will contribute for new insights into the innate immune response in leprosy immunoregulation and may indicate potential new targets for therapeutic intervention.

## Data Availability

The dataset supporting the findings of this article is available from the corresponding author upon request.

## Abreviations

MBL: mannose binding-lectin
SNV: single nucleotide variant
PCR-RFLP: polymerase chain reaction-restriction fragment length polymorphism
MBP-oligomer: oligomeric MBL carbohydrate-binding domain
ELISA: enzyme linked immunosorbent assay
OR: odd ratio
CI: confidence interval
M. leprae: Mycobacterium leprae
WHO: world health organization
L: leprosy patients
HC: healthy controls
MB: multibacillary patients
PB: paucibacillary patients
LL: lepromatous leprosy
DL: dimorph leprosy
IDL: indeterminate leprosy
TL: tuberculoid leprosy
TMB: tetramethylbenzidine
DNA: deoxyribonucleic acid.

## Ethics approval and consent to participate

The study protocol was approved by The Local Ethic Medical Committee (CAEE n^o^. 19679119.8.0000.5244). Written informed consents were obtained from the participants and/or yours parents/guardians of the participating children.

## Consent for publication

Not applicable.

## Authors’ contributions

EMS, acquisition of data and drafting of manuscript; LMM, analysis and interpretation of data; RCME, acquisition of data; BVGF, acquisition of data; YECM, acquisition of data; TLK, In memoriam; EPNJr, clinical examination of patients; WDS, analysis of data; ALPR, study conception and design, interpretation of data, and drafting of manuscript.

## Acknowledgements

We thank Hansen Health Program of the Campos dos Goytacazes Health Secretariat and the dermatologist Edilbert P. Nahn Jr. for their hard work and technical expertise in selecting patients and collecting biological material. We also thank the local Blood Bank from Ferreira Machado Hospital, Campos dos Goytacazes medical staff, in selecting healthy controls and collecting biological material.

